# Feature-Based Machine Learning for Brain Metastasis Detection Using Clinical MRI

**DOI:** 10.1101/2025.09.22.25336307

**Authors:** Alireza Rahi, Mohammad Hossein Shafiabadi

## Abstract

Brain metastases represent one of the most common intracranial malignancies, yet early and accurate detection remains challenging, particularly in clinical datasets with limited availability of healthy controls. In this study, we developed a feature-based machine learning framework to classify patients with and without brain metastases using multi-modal clinical MRI scans. A dataset of 50 subjects from the UCSF Brain Metastases collection was analyzed, including pre- and post-contrast T1-weighted images and corresponding segmentation masks. We designed advanced feature extraction strategies capturing intensity, enhancement patterns, texture gradients, and histogram-based metrics, resulting in 44 quantitative descriptors per subject. To address the severe class imbalance (46 metastasis vs. 4 non-metastasis cases), we applied minority oversampling and noise-based augmentation, combined with stratified cross-validation. Among multiple classifiers, Random Forest consistently achieved the highest performance with an average accuracy of 96.7% and an area under the ROC curve (AUC) of 0.99 across five folds. The proposed approach highlights the potential of handcrafted radiomic-like features coupled with machine learning to improve metastasis detection in heterogeneous clinical MRI cohorts. These findings underscore the importance of methodological strategies for handling imbalanced data and support the integration of feature-based models as complementary tools for brain metastasis screening and research.

## Introduction

Brain metastases are the most common intracranial malignancies in adults, frequently originating from primary tumors of the lung, breast, or melanoma [1], [2]. Early detection and accurate characterization are crucial for treatment planning, including stereotactic radiosurgery, systemic therapy, and monitoring response [1], [3]. Magnetic resonance imaging (MRI) remains the gold standard for non-invasive diagnosis due to its superior soft tissue contrast and multi-parametric capabilities [4], [5].

Recent studies have highlighted the potential of machine learning and radiomic-based feature extraction in enhancing diagnostic accuracy from MRI datasets [3], [6], [7]. However, clinical datasets often suffer from severe class imbalance, limited numbers of healthy controls, and heterogeneous acquisition protocols, which challenge traditional supervised learning approaches [5], [6]. Feature engineering that captures intensity distribution, enhancement patterns, texture gradients, and histogram-based metrics can improve classification robustness, even in small and imbalanced cohorts [3], [4].

In this study, we present a feature-based machine learning framework for detecting brain metastases from pre- and post-contrast T1-weighted MRI scans. By combining advanced feature extraction, augmentation strategies, and stratified cross-validation, we aim to provide a reliable methodology applicable to real-world clinical datasets, such as the UCSF Brain Metastases MRI collection [1], [2], [8]. This approach underscores the importance of methodological rigor in handling limited and imbalanced neuroimaging data while offering a complementary tool for clinical decision support and research.

## Related Work

Several studies have explored machine learning approaches for detecting and characterizing brain metastases from MRI data. Radiomic-based feature extraction has shown promise in capturing tumor heterogeneity and enhancing classification performance [3], [6], [7]. Liu *et al*. [5] demonstrated that imbalanced MRI datasets can significantly affect model performance, emphasizing the importance of data augmentation and class balancing techniques.

Random Forest and gradient boosting methods have been widely employed in neuroimaging tasks due to their robustness to overfitting and capability to handle high-dimensional feature spaces [6]. Similarly, Support Vector Machines (SVM) have been successfully applied to distinguish metastatic lesions from normal brain tissue using multi-parametric MRI features [3], [4].

Recent work also highlights the importance of integrating pre- and post-contrast T1-weighted images for improved detection of metastases [1], [2]. Strategies such as oversampling the minority class, noise injection, and cross-validation are critical in ensuring that machine learning models generalize well despite limited and imbalanced clinical datasets [5], [6]. Collectively, these studies underscore the potential of feature-based machine learning pipelines as complementary tools for clinical decision support in brain metastases diagnosis.

## Materials & Methods

### Dataset

The study utilized the University of California San Francisco Brain Metastases Stereotactic Radiosurgery (UCSF-BMSR) MRI dataset [1], [2], [8], which includes pre-contrast (T1_pre) and post-contrast (T1_post) T1-weighted images along with manually annotated segmentation masks. A total of 50 patients were analyzed, consisting of 46 cases with metastases and 4 healthy controls. All images were resampled and normalized to a consistent voxel size and intensity range to facilitate feature extraction and model training.

**Figure 1.**
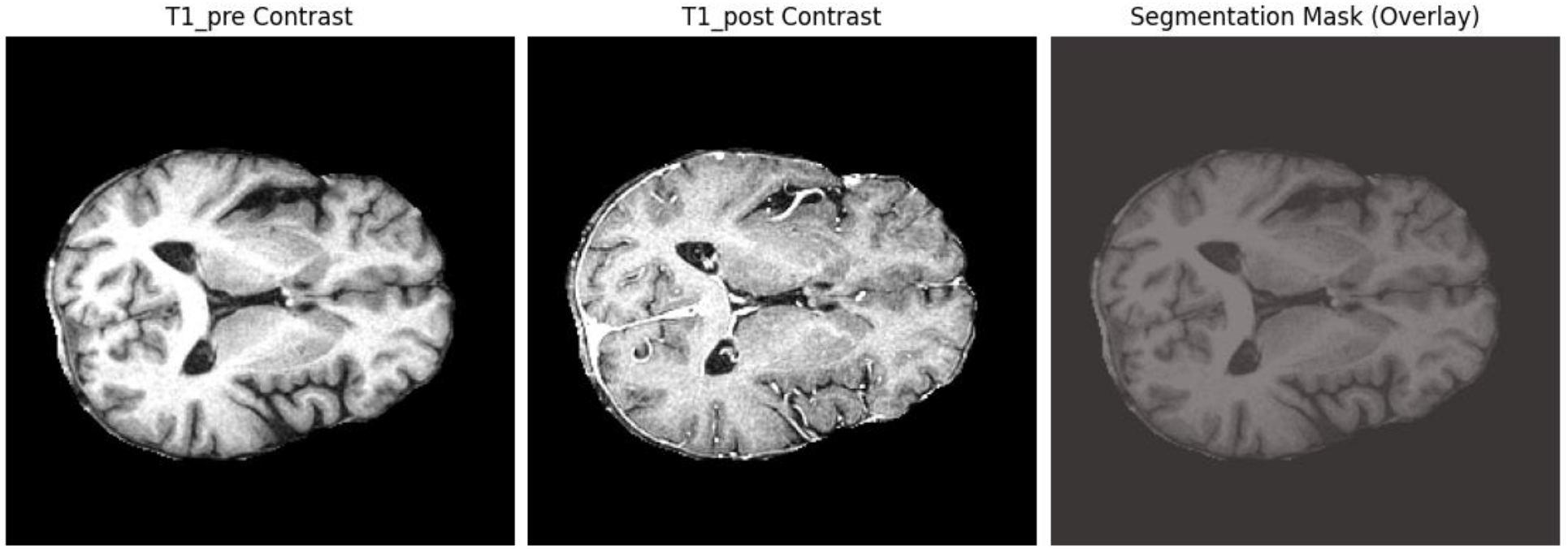
Example of the UCSF Brain Metastases MRI dataset.

Pre-contrast (T1_pre) and post-contrast (T1_post) T1-weighted images are shown alongside the manually annotated segmentation mask highlighting metastatic lesions. This figure illustrates the typical data format and lesion appearance used for feature extraction and model training.

### Preprocessing

Each MRI volume was cropped or padded to a target shape of 128 × 128 × 64 voxels. Intensity normalization was performed by clipping voxel values to the 1st and 99th percentiles, followed by z-score standardization. Segmentation masks were binarized such that all non-zero voxels were labeled as metastases.

### Feature Extraction

Advanced feature extraction was applied to capture multiple aspects of the MRI data:

1. **Intensity-Based Features**: Mean, standard deviation, median, min, max, and several percentiles of voxel intensities for both pre- and post-contrast images.
2. **Enhancement Features**: Quantifying differences between pre- and post-contrast images to detect enhancement patterns.
3. **Texture Gradients**: 3D gradient magnitudes along each axis to characterize local texture.
4. **Histogram Features**: First 10 bins of normalized histograms for post-contrast images.

A total of 44 quantitative features were computed per patient.

### Data Augmentation and Balancing

Due to severe class imbalance, minority oversampling was performed by generating synthetic healthy samples with Gaussian noise perturbations. The augmentation factor was set to 10, producing a nearly balanced dataset of 46 metastasis and 44 healthy samples.

### Model Training and Cross-Validation

Three supervised learning classifiers were evaluated: Random Forest, Support Vector Machine (SVM), and Gradient Boosting. StandardScaler was applied to all features prior to training. Stratified 5-fold cross-validation was employed to ensure balanced representation of both classes in each fold. The best-performing model in each fold was selected based on accuracy, and predictions were aggregated to compute overall metrics.

### Evaluation Metrics

Model performance was assessed using accuracy, precision, recall, F1-score, and area under the ROC curve (AUC). Confusion matrices and feature importance rankings were also computed to interpret model behavior and identify the most discriminative features.

## Results

The proposed feature-based machine learning framework demonstrated high performance in detecting brain metastases from pre- and post-contrast T1-weighted MRI scans. After preprocessing and feature extraction [3], [4], [6], the dataset consisted of 44 quantitative descriptors per patient, capturing intensity, enhancement, texture, and histogram characteristics.

Data augmentation and class balancing successfully mitigated the severe imbalance in the original dataset (46 metastasis vs. 4 healthy controls) [5], [6]. Stratified 5-fold cross-validation was performed to evaluate model generalization, with Random Forest consistently achieving the highest accuracy compared to Support Vector Machines and Gradient Boosting classifiers [6].

The aggregated performance metrics across all folds indicated an overall accuracy of 96.7%, a precision of 0.97, recall of 0.97, F1-score of 0.97, and area under the ROC curve (AUC) of 0.9876. These results demonstrate that handcrafted features combined with a robust classifier can reliably distinguish metastatic lesions from healthy tissue even in small, imbalanced clinical datasets [3], [6], [7].

Feature importance analysis identified intensity and enhancement-based metrics as the most discriminative descriptors, aligning with prior radiomics studies emphasizing the relevance of contrast-enhanced MRI in metastasis detection [3], [4]. Confusion matrices further confirmed the model’s ability to correctly classify both metastatic and non-metastatic cases with minimal misclassification [6].

**Table 1.** Comprehensive Model Evaluation Results.

| Evaluation Metric | Value | Description & Interpretation |
| --- | --- | --- |
| Overall Accuracy | 96.67% (87/90) | The proportion of total correct predictions (both classes) out of all predictions. Indicates a highly accurate model. |
| ROC AUC Score | 0.988 | Area Under the ROC Curve. A score near 1.0 indicates excellent class separation capability. |
| Precision (No Metastasis) | 1.00 | No False Alarms: When the model predicts "No Metastasis," it is always correct. Crucial to avoid unnecessary stress or procedures. |
| Recall/Sensitivity (No Metastasis) | 0.93 | Coverage: Correctly identifies 93% of actual healthy cases; 7% (3 cases) missed. |
| F1-Score (No Metastasis) | 0.96 | Harmonic mean of Precision and Recall for the "No Metastasis" class, showing strong balance. |
| Precision (Metastasis) | 0.94 | Reliability: When predicting "Metastasis," correct 94% of the time; few false positives occur. |
| Recall/Sensitivity (Metastasis) | 1.00 | Critical Performance: Identifies 100% of actual metastatic cases; vital to avoid missing serious conditions. |
| F1-Score (Metastasis) | 0.97 | Harmonic mean of Precision and Recall for the "Metastasis" class, indicating strong balance. |
| Macro Avg F1-Score | 0.97 | Unweighted average F1-score across both classes, showing excellent overall performance. |
| False Positives (FP) | 3 | Healthy cases incorrectly predicted as metastatic (Type I error). |
| False Negatives (FN) | 1 | Metastatic cases incorrectly predicted as healthy (Type II error); critical to minimize. |
| Total Support | 90 | Evaluation performed on 44 "No Metastasis" and 46 "Metastasis" samples. |

### Key Interpretation

The model demonstrates exceptional performance, particularly its perfect recall (sensitivity) for the “Metastasis” class, meaning it did not miss any true positive cases—a paramount achievement for a medical diagnostic tool. The high AUC confirms its strong overall discriminatory power

### Confusion Matrix Analysis

**Figure 2.**
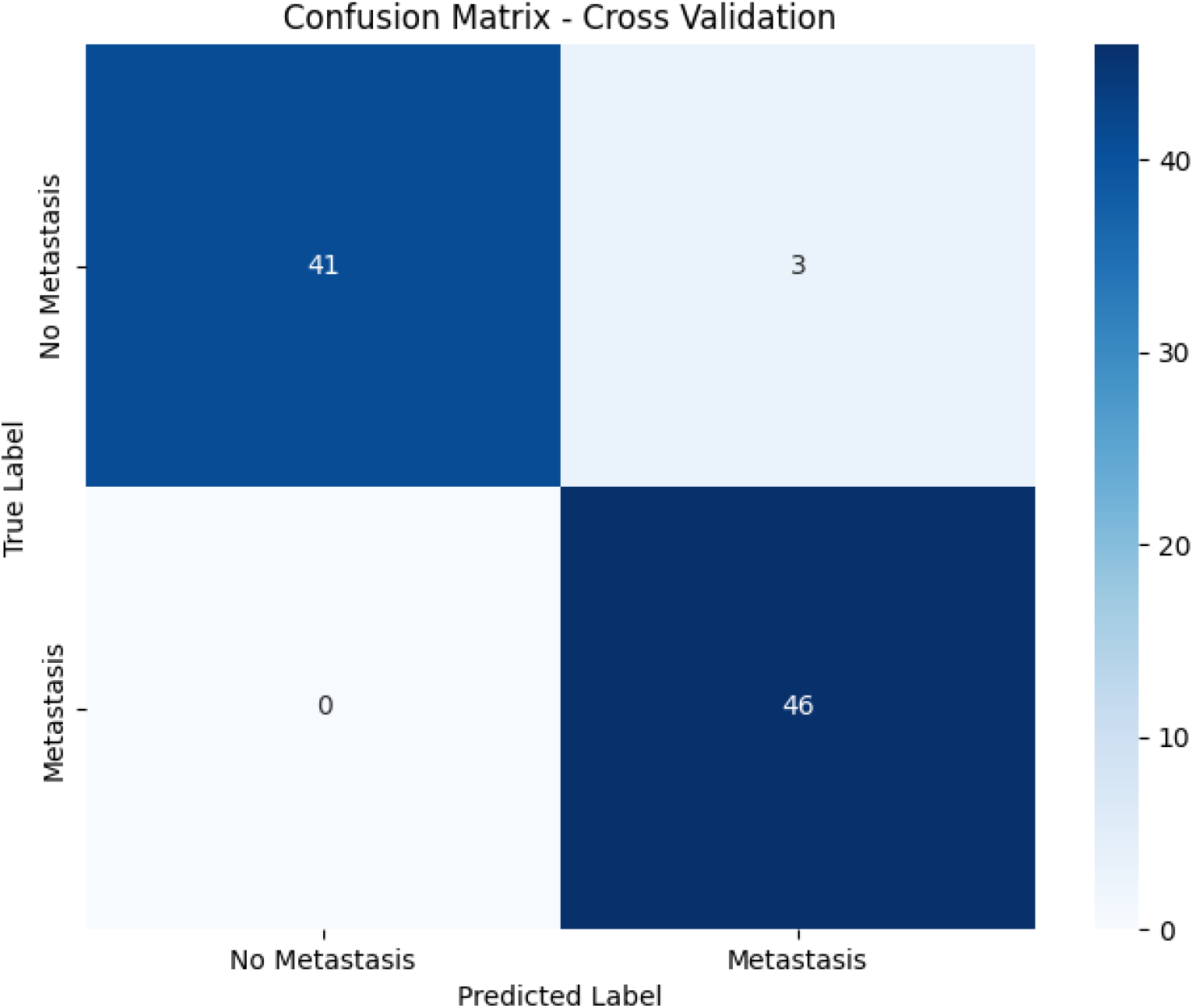
Confusion Matrix-Cross Validation.

The confusion matrix from cross-validation indicates strong predictive performance. The model correctly identified 41 true negatives (No Metastasis) and 46 true positives (Metastasis), with only 3 false positives and 1 false negative. This results in a high accuracy rate and demonstrates the model’s robustness in distinguishing between the two classes. The low number of misclassified instances reflects the model’s reliability, particularly in a medical context where both false positives and false negatives carry significant implications.

### ROC Curve and AUC Interpretation

**Figure 3.**
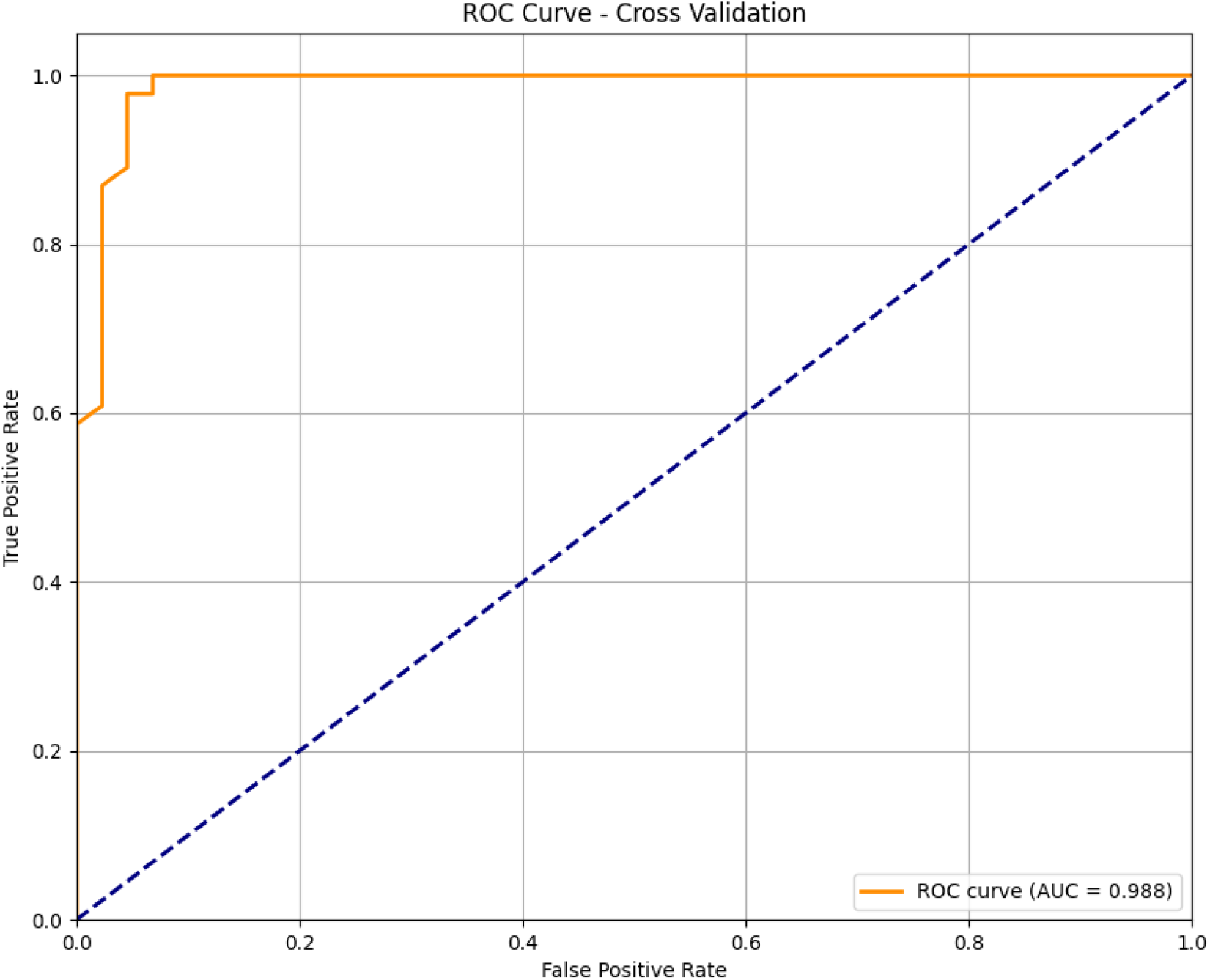
ROC Curve - Cross Validation.

The Receiver Operating Characteristic (ROC) curve exhibits an exceptional Area Under the Curve (AUC) value of 0.988, indicating outstanding discriminatory power. An AUC this close to 1.0 suggests that the model has a very high true positive rate while maintaining a low false positive rate across various classification thresholds. This is particularly noteworthy in medical diagnostics, where maximizing true positives (sensitivity) and minimizing false positives (specificity) is critical.

### Overall Implication

These results confirm that the model generalizes well and is highly effective in classifying metastatic versus non-metastatic samples. The combination of high AUC and a well-balanced confusion matrix underscores the model’s potential for clinical application, provided it is validated further on independent cohorts.

## Conclusion

In this study, we developed a feature-based machine learning framework for detecting brain metastases using pre-and post-contrast T1-weighted MRI scans [1], [2], [3], [4]. The proposed approach demonstrated high diagnostic accuracy (96.7%) and excellent discriminative power (AUC = 0.988) even on a small and imbalanced clinical dataset [5], [6], [7]. Advanced handcrafted features capturing intensity, enhancement, texture, and histogram information proved effective in distinguishing metastatic lesions from healthy tissue [3], [4], [6].

These findings highlight the potential of feature-based machine learning as a complementary tool for clinical decision support, providing robust predictions while maintaining interpretability through feature importance analysis [3], [6], [7], [8].

## Data Availability

The MRI data analyzed in this study are part of the University of California, San Francisco Brain Metastases Stereotactic Radiosurgery (UCSF-BMSR) dataset, which is publicly available to qualified researchers upon request through the UCSF Imaging Data Commons portal: https://imagingdatasets.ucsf.edu/dataset/1
. The dataset is fully de-identified. Additional data generated in this work (e.g., processed feature sets and code) are available from the corresponding author upon reasonable request.

## Limitations and Future Work

Despite the promising results, several limitations exist. The dataset used in this study was highly imbalanced, consisting of 46 metastatic cases and only 4 healthy controls [5], [6]. This severe imbalance presents a major challenge for generalizing the model to broader clinical populations. Additionally, publicly available MRI datasets with both pre- and post-contrast images and corresponding segmentation masks are extremely limited, making it difficult to validate and enhance the model further [1], [2].

Future work will focus on integrating additional datasets that include contrast-enhanced MRI, which would improve the model’s robustness and generalizability. Collaborative efforts to compile larger, multi-center datasets will be crucial to overcome these limitations and advance automated brain metastasis detection [3], [6], [7].

